# Implementation of SARS-CoV2 Screening in K-12 Schools using In-School Pooled Molecular Testing and Deconvolution by Rapid Antigen Test

**DOI:** 10.1101/2021.05.03.21256560

**Authors:** Nira R. Pollock, David Berlin, Sandra C. Smole, Lawrence C. Madoff, Kelsey Henderson, Elizabeth Larsen, Jeremiah Hay, Stacey Gabriel, Atul A. Gawande, Niall J. Lennon

## Abstract

**What is already known about this topic?:** SARS-CoV2 testing is a key component of a multi-layered mitigation strategy to enable safe return to in-person school for the K-12 population. However, costs, logistics, and uncertainty about effectiveness are potential barriers to implementation.

**What is added by this report?:** Over three months, 259,726 individual swabs were tested across 50,636 pools from 582 schools. Pool positivity rate was 0.8%; 98.1% of pools tested negative and 0.3% inconclusive, and 0.8% of pools submitted could not be tested. In reflex testing, 92.5% of fully deconvoluted pools with N1 or N2 target Ct ≤30 yielded a positive individual using the BinaxNOW antigen rapid diagnostic test (Ag RDT) performed 1-3 days later. With sufficient staffing support and low pool positivity rates, pooled sample collection and reflex testing were feasible for schools.

**What are the implications for public health practice?:** Screening testing for K-12 students and staff is achievable at scale and at low cost with a scheme that incorporates in-school pooling, RT-PCR primary testing, and Ag RDT reflex/deconvolution testing. Staffing support is a key factor for program success.

---

In March of 2021, the U.S. Department of Health and Human Services announced an investment of $10 billion under the American Rescue Plan to increase screening testing to help schools reopen (*1*). Pooled testing, a form of group test whereby more than one individual specimen is combined prior to the laboratory test process, can provide affordable testing at this scale. Many scientists and epidemiologists have advocated for pooled testing for the detection of the SARS-CoV2 virus in large population cohorts (*2–5*). Several strategies for pooled testing have been proposed. The simplest pooling design dates back to the work of Robert Dorfman in 1943 (*6*). If the pool test result is negative, the members of the pool are presumed to be negative. If the pool test result is positive, the constituent members of the pool must be tested individually (known as “pool deconvolution,” or “reflex testing”) to determine who is actually positive for the test analyte.

The Broad Institute has established a distributed Dorfman pooled testing process, termed “Pod Pooling,” in which the pooling event happens at the site of specimen collection. In this model, individual anterior nasal (AN) swab samples are placed into a single dry sample tube (no transport media) with a maximum of 10 swabs per tube. At the laboratory, highly automated processes are employed for accessioning, decapping, swab rehydration, sample transfer, RNA extraction, and target analyte detection. The downstream testing for pooled specimens (from extraction onwards) follows exactly the same process as the laboratory’s EUA clinical diagnostic RT-PCR test for SARS-CoV2 (https://www.fda.gov/media/146499/download).

In order for Pod Pooling to be effective, four things must be in place: 1) short turnaround times for pooled test results; 2) pooled test capacity; 3) rapid reflex testing; and a robust result reporting system. The testing platform routinely delivers results within 12-16 hours from sample receipt. This translates into a median end-to-end turnaround time experienced by the sites of 20.7 hours. The testing laboratory operates 24/7 and has a current capacity for pooled tests of 40,000 per day (equating to up to 400,000 individuals per day). The requirement for collection of a new sample for pool deconvolution in this program design led to the idea of using antigen (Ag) rapid diagnostic tests (RDTs) for reflex testing, leveraging a recent evaluation of the performance of the Abbott BinaxNOW COVID-19 Ag Card (*7*) in adults and children (*8*) and state procurement of this test at the scale required for this use case. On January 8th, 2021, the Massachusetts Departments of Early and Secondary Education and Public Health announced a new asymptomatic pooled testing program utilizing this design for MA schools (*9*). In order to facilitate school participation in the program, MA health officials distributed sample standing orders for school or other local providers and added the participating schools as locations under the State Public Health Laboratory’s CLIA Certificate of Waiver.

At the pooling site, asymptomatic students and staff are organized into pools of two to ten members. Observed self-collection of AN swabs is employed for adults and older students (grades 2-12), and younger students are swabbed by designated staff. Schools keep track of which individuals are in which pool, either on paper or with the help of provided software. All the swabs in a pool are tested together and the lab reports a single group result to the registrant. If a pool test returns positive, every individual in the pool must temporarily take precautions as though they are positive for COVID and follow-up with an individual (reflex) test. Program instructions guided schools to utilize BinaxNOW (with AN swab collected by trained staff) for positive pool deconvolution; if all individuals tested negative with BinaxNOW, schools could choose between performing reflex PCR or a second round of BinaxNOW. Deidentified pool deconvolution data provided voluntarily by schools working with one operations partner (CIC Health) were available for analysis, allowing assessment of the performance of BinaxNOW in this testing scenario.

Between Jan 2021 and April 2021, pooled AN swab RT-PCR testing with Ag RDT reflex testing was implemented at schools in Massachusetts, including 582 schools submitting testing to the Broad laboratory and additional schools submitting to another laboratory (Ginkgo Bioworks). During this period, 259,726 individuals were tested across 50,636 pools submitted by schools to Broad with assistance from operational partners CIC Health and Project Beacon. The average number of swabs per pool was 7 (range 2-10). The positive pool rate was 0.8%. The median turnaround time from pooled swab collection to result return was 20.7 hours (see Table 1).

**Table 1.** High level metrics of the school testing program as of April 9th, 2021. Numbers are for schools included in the Massachusetts pilot program that were processed for testing at the Clinical Research Sequencing Platform (CRSP) at the Broad Institute.

| **Metric** | **Number** |
| --- | --- |
| Number of schools sending pools so far | 582 |
| Number of pooled tests run | 50,636 |
| Median number of swabs per pool | 7 |
| Range of number of swabs per pool | 2-10 |
| Number of individuals tested in pools<sup>1</sup> | 259,726 |
| Median TAT(hrs) from collection time to result return | 20.7 |
| Interquartile range of TAT (hrs) | 14 - 28.4 |

TAT, turn-around time. <sup>1</sup>Total number of swabs; note that individuals may be tested in more than one pool over time (i.e. repeat testing).

The mean RT-PCR cycle threshold (Ct) values for positive pools were 26.1 and 27.6 for the N1 and N2 viral targets, respectively (see Table 2), equating to a mean viral load of 9.3 x 10^4^ copies/mL based on the assay standard curve. Assay validation data had demonstrated that positive pool Cts are ∼1.6 Ct greater than the individual swab Ct, an expected finding based on the fixed dilution factor used for pools (5mL of rehydration buffer added to tube regardless of number of swabs) compared to individual swab testing (1mL added to one swab) (*10*). Pollock *et al* (*8*), in a large study comparing BinaxNow and Broad RT-PCR tests performed on paired AN swabs from symptomatic and asymptomatic adults and children, reported 95.8% sensitivity of the BinaxNOW (all subgroups combined) when the RT-PCR Ct was ≤30, and 81.2% sensitivity with Ct ≤35. Based on the dilution factor Ct shift, 76.6% (321/419) of the positive pools would be predicted to have an individual swab with N1 or N2 target Ct≤30 and 99.0% (415/419) of pools a sample with Ct ≤35. However, the impact of the time interval between pooled sample collection and deconvolution testing (expected to range from 1-3 days) on viral load--and thus BinaxNOW sensitivity--was unknown.

**Table 2.** Pooled testing results as of April 9th, 2021. Results are based on a RT-PCR test with multiplexed N1, N2, and RP targets as outlined in the assay EUA (EUA200147).

| **Metric** |  | **Number** | **%** |
| --- | --- | --- | --- |
| **Number of pooled tests run** |  | **50,636** | **100** |
| Number of negative pools<sup>a</sup> |  | 49,663 | 98.1 |
| Number of positive pools<sup>b</sup> |  | 419 | 0.8 |
| Number of invalid pools<sup>c</sup> |  | 41 | 0.1 |
| Number of inconclusive pools<sup>d</sup> |  | 88 | 0.2 |
| Unsatisfactory Specimens (test not performed) |  |  |  |
|  | Reason - at least one swab upside down in tube | 288 | 0.6 |
|  | Reason - sample received >56hrs after collection | 73 | 0.1 |
|  | Reason - lab incident | 53 | 0.1 |
|  | Reason - test ordered but no sample sent | 5 | 0.01 |
|  | Reason - sample too viscous after rehydration | 4 | 0.01 |
|  | Reason - tube label data does not match order | 2 | 0.004 |
| Positive pool Viral N1 target Ct - Mean (std dev) |  | 26.1 (5.6) |  |
| Positive pool Viral N1 target Ct - Range |  | 15 - 37.8 |  |
| Positive pool Viral N2 target Ct - Mean (std dev) |  | 27.6 (6.3) |  |
| Positive pool Viral N2 target Ct - Range |  | 15.3 - 39.8 |  |

a, negative pools have no detected viral targets and a positive human control target (RP gene);
b, Positive pools have two detected viral targets (N1 and N2); c, Invalid pools have no detected viral targets and no detected human control target; d, Inconclusive pools have only one viral target detected. Ct, cycle threshold.

Between 1/4/21 and 4/9/21, 124 positive school pools were followed by BinaxNOW testing for deconvolution and had both pooled PCR Ct data and reflex results available for analysis. In 10/124 pools, not all individuals in the pool were tested by BinaxNOW (due to factors including electing to do PCR elsewhere, development of symptoms, refusal, or quarantine). Deconvolution results for the 114 pools with full BinaxNOW deconvolution are presented in Table 3. For each of the five deconvolution outcomes, the % of pools with either N1 or N2 Ct value ≤30 is shown. Of 80 fully deconvoluted pools with either N1 or N2 Ct value ≤30, 74 (92.5%) yielded an individual with a positive BinaxNOW result. BinaxNOW was performed 1-2D after PCR testing in 93.0% of pools, and within 1D in 75.4% (range 1-3D).

**Table 3.** Reflex testing performance.

| **Metric** | **Number (%)** | **Pool N1 or N2 Ct ≤ 30**<br>**n (%)** |
| --- | --- | --- |
| Positive pools with available reflex test data | 124 | 86/124 (69.4) |
| Pools with complete BinaxNOW deconvolution (all individuals tested by BinaxNOW) | 114/124 (91.9) | 80/114 (70.2) |
| True positive BinaxNOW<sup>a</sup> | 89/114 (78.1) | 74/89 (83.1) |
| False negative BinaxNOW<sup>b</sup> | 5/114 (4.4) | 1/5 (20.0) |
| Likely false negative BinaxNOW<sup>c</sup> | 4/114 (3.5) | 1/4 (25.0) |
| Pool PCR result unconfirmed<sup>d</sup> | 12/114 (10.5) | 2/12 (16.7) |
| Unknown<sup>e</sup> | 4/114 (3.5) | 2/4 (50.0) |

a, individual in pool tested positive by BinaxNOW (in 88/89 cases, only one positive BinaxNOW result was observed; in 1/89, two were observed (the two individuals were from the same family)); b, all BinaxNOW results were negative, but an individual in the pool tested positive by reflex PCR; c, all BinaxNOW results were negative, but an individual in the pool had an inconclusive reflex PCR result (only 1 of 2 PCR targets positive); d, all BinaxNOW and additional reflex testing results (repeat BinaxNOW testing (n = 4) or PCR testing (n = 8) on all individuals in the pool) were negative, making it impossible to confirm the positive pool PCR result; e, all BinaxNOW results were negative, but additional reflex PCR results were not available. Ct, cycle threshold; PCR, polymerase chain reaction

An informal survey of school districts participating in the testing program identified a consistent need for additional staff support in order to successfully implement the program, given time and personnel requirements for in-school pool collection, communication with individuals in positive pools, and reflex testing.

## Discussion

The three months of Pod Pooling data indicate that this method can be successfully implemented in the school setting. The distributed pooling model facilitates scale and rapid turnaround and keeps laboratory costs low (latter in contrast to some strategies in which sample pooling and reflex testing are performed by the laboratory). The low rate of unsatisfactory incoming samples indicates that users in the school setting can effectively follow provided testing protocols. It should be noted that implementation of this model requires attention to staffing needs for in-school sample collection and reflex testing, and a commitment from participants to return for reflex testing, which can be challenging for some families and staff.

This pooled testing model requires a feasible and reliable reflex testing strategy that can be deployed as soon as possible after a positive pool is detected. The use of an Ag RDT significantly shortens the period of time to generate (and act on) an individual level result.

BinaxNOW pool deconvolution was successful in 92.5% of pools with Ct values ≤30 for either N1, N2, or both targets, which represents 73.3% (307/419) of positive pools submitted in the program to date; pools with low Ct values are expected to correspond to individuals with higher infectivity. BinaxNOW was performed 1-2 D after PCR testing in 93% of pools. Only 9 confirmed false negative or likely false negative BinaxNOW results were observed out of 114 fully completed deconvolution attempts, and of those, 7 (77.8%) had pool Ct >30 for both targets. However, given that 15/89 pools (16.9%) with positive BinaxNOW deconvolution results had Ct >30 for both N1 and N2, and given the benefit of rapid case identification, it may be worthwhile to attempt BinaxNOW regardless of Ct value of the positive pool. Of note, interpretation of positive BinaxNOW reflex results as true positives was felt to be justified by the context (positive pool) and the high observed specificity of BinaxNOW (*8*).

It should be noted that 25.8% of BinaxNOW pool deconvolution testing attempts did not detect a positive individual, requiring either reflex PCR done within the school program or, in some cases, outside reflex PCR testing for individuals who did not return for BinaxNOW testing. These early program data help to highlight the logistic complexity of requiring a return visit for reflex testing after a positive pool, and the necessity of having a plan for expedited reflex PCR available for all participants. Thus far, performing BinaxNOW tests on two consecutive days if the first round of BinaxNOW testing is negative has not yielded any positive BinaxNOW results. Analysis of reflex results for pools with inconclusive pool PCR results is underway. Finally, optimization to streamline reflex testing procedures, particularly for large districts with student/staff transportation and communication challenges, is in progress.

## Data Availability

The work here represents summary data from the implementation of testing. No individual level patient data is made available as part of this work.

## Acknowledgements

We acknowledge the work of Massachusetts school districts to implement the asymptomatic pooled testing program described in this manuscript, and the efforts of the Massachusetts Safer Teachers, Safer Students Testing Collaborative to gather operational data to inform program optimization. We thank Dr. Al DeMaria for his comments on the manuscript. The work at the Broad Institute was funded in part by the NIBIB RADx Advanced Technology Program and the Commonwealth of Massachusetts.

## Funding Statement

This project was supported by the Centers for Disease Control and Prevention’s Coronavirus Aid, Relief, and Economic Security (CARES) Act of 2020 within Project E: Emerging Infections ELC Reopening Schools (Grant #6 NU50CK000518-02-06) of the U.S. Department of Health and Human Services (HHS) as part of a financial assistance award totaling $205M. The contents are those of the author(s) and do not necessarily represent the official views of, nor an endorsement by, CDC/HHS or the U.S. Government.

The work at the Broad Institute was funded, in part, by the NIBIB RADx Advanced Technology Program and the Commonwealth of Massachusetts.

## Notes

### Competing Interest Statement

The authors have declared no competing interest.

### Funding Statement

This project was supported by the Centers for Disease Control and Prevention Coronavirus Aid, Relief, and Economic Security (CARES) Act of 2020 within Project E: Emerging Infections ELC Reopening Schools (Grant #6 NU50CK000518-02-06) of the U.S. Department of Health and Human Services (HHS) as part of a financial assistance award totaling $205M. The contents are those of the author(s) and do not necessarily represent the official views of, nor an endorsement by, CDC/HHS or the U.S. Government.
The work at the Broad Institute was funded, in part, by the NIBIB RADx Advanced Technology Program and the Commonwealth of Massachusetts.

### Author Declarations

Massachusetts Department of Public Health IRB reviewed and approved the work (and considered it not human subjects research).

